# “*Why are you doing contraception? … They don’t need it in jail”.* Service provider views on women’s access to contraception in Australian prisons

**DOI:** 10.1101/2025.06.11.25329445

**Authors:** Helene Smith, Mandy Wilson, Basil Donovan, Jocelyn Jones, Tony Butler, Paul Simpson

**Affiliations:** School of Population Health, University of New South Wales, Sydney, Australia; Kirby Institute, University of New South Wales, Sydney, Australia; Kurongkurl Katitjin, Centre for Indigenous Australian Education & Research, Edith Cowan University, Perth, Australia; National Drug & Research Institute, Curtin University, Perth, Australia

## Abstract

**Introduction:** Women with a history of incarceration report higher rates of pregnancy, pregnancy terminations, and the use of Long-Acting Reversible Contraception (LARC) methods, alongside poorer birth outcomes compared to non-justice involved women, suggesting distinctive family planning needs. Despite this, little is known on how this is provided within carceral settings, or the structural barriers experiences by both staff and other service providers. In this article, we report findings from a qualitative study conducted with a sample of professionals working in New South Wales (NSW), Australia, who either provide healthcare services to women either during incarceration or in the immediate post-release period. Specifically, we describe structural factors that may impede or support access to the provision, insertion or removal of contraception devices.

**Methods:** We conducted semi-structured interviews with n=12 professionals, working in NSW, who either provide reproductive healthcare services to women during incarceration or in the immediate post-release period. Data was analysed thematically against the Vulnerable Populations Behaviour Model.

**Results:** Seven themes were identified relating to contraception service provision in prison. Participants observed specific patient characteristics which they believed were driving contraception needs including 1) a state of interrupted fertility caused by incarceration, and 2) post-release chaos which impeded access to appropriate family planning in the community. Areas of high need were characterised by 1) accessible, reversible and non-invasive contraception options, and 2) thoughtful counselling. Structural factors impeding contraception service access during incarceration included: 1) service priorities and staff biases negatively impacting access, 2) poor overall health service integration, and 3) inequitable funding models during incarceration. Consequently, incarcerated women do not have comparable access to contraception compared to women in the community.

**Conclusion:** The lack of focus on contraception services in prison represents a missed opportunity to support highly marginalised women who may also face challenges accessing such services upon release.

## Introduction

Australian women with a history of incarceration have distinctive contraception-use profiles and pregnancy histories compared to women in the community[1–6]. This includes higher rates of pregnancies, pregnancy terminations[1–3], lower rates overall of contraceptive use[4] but higher rates of Long-Acting Reversible Contraceptive (LARC) methods[3], as well as poorer birth outcomes[5, 6].

The provision of family planning services, including access to appropriate and reversible modern contraception, has long been regarded as a crucial service in ensuring economic empowerment for women along with reducing maternal and child morbidity and mortality[7, 8]. It is increasingly recognised that for many women, pregnancy planning and intention can be fluid concepts[9–15] and critical human rights and Reproductive Justice principles need to be prioritised when these services are provided to marginalised women.

Human rights considerations include the provision of accessible and non-judgmental services, quality counselling, informed decision-making and non-coercive practices, particularly in relation to implants and intrauterine devices (IUDs)[16–18]. Reproductive Justice advocates go further to recognise that women’s ability to enact their true reproductive choices are often hampered by intersecting and systematic oppressions. These can be driven by legal and institutional structures, as well as demographic factors such as race or socio-economic status [19–22]. These issues are further compounded within a carceral setting where questions of coercion can become highly precarious[23, 24].

Questions of Reproductive Justice have been examined in Australia in the context of how to provide culturally safe sexual and reproductive health (SRH) services to Indigenous women and girls[25, 26], abortion law reform[27–29] and accessing family planning services during COVID-19[30]. These studies however have not considered the implications within a carceral setting, which is over-represented by women from marginalised communities, including a large proportion of Aboriginal and Torres Strait Islander women[31]. Many women with a history of incarceration are more likely to have experienced significant trauma[1] and substance abuse histories[32], compared to women in the community. In addition, carceral settings often report unique barriers to accessing healthcare services in general including long-wait times[33–35], administrative disturbances[36], judgmental and racist custodial staff[33, 35], distrust of authority figures[37] and low levels of awareness by incarcerated persons of what health services are available[33].

When considering challenges in the provision of contraception in the general community, particularly LARC methods, qualitative studies with healthcare workers in Australia have highlighted several barriers[38–43]. These include limited training and confidence in performing LARC insertions[38–40], patient misconceptions around side-effects[38, 40], provider preferences dominating patient preferences[41, 42], provider discomfort with discussing sexual wellbeing[41, 42], conflicting views on the appropriateness of LARC removals[43] and inadequate reimbursement models for providers[39]. However, no studies have considered whether these issues are mirrored in the Australian carceral system, a healthcare model which is state funded and unable to access federally funded schemes such as Medicare or the Pharmaceutical Benefits Scheme[44, 45]. Both schemes, which provide free or highly subsidised health services and medicines, are key mechanisms for the provision of accessible contraception in Australia including LARC insertions and removals, General Practitioner (GP) consultations to support both counselling and the prescription of oral contraception.

In this article, we report findings from a qualitative study conducted with a sample of professionals working in New South Wales (NSW), who either provide reproductive healthcare services to women during incarceration or in the immediate post-release period. Specifically, we describe structural factors that may impede or support access to the provision, insertion or removal of contraception devices.

## Methods

### Study Design

Between August 2022 and July 2023, a qualitative study was conducted, where 12 participants were recruited to complete a one-hour semi-structured interview with one of the authors (HS).

Both interview guides and the coding framework were informed by the Behavioural Model for Vulnerable Populations[46], which offers an empirically supported framework to understand the combination of multi-level factors, termed “predisposing characteristics” (that is, demographic and social structure characteristics of individuals accessing the service), “need factors” (perceptions and evaluated needs of special relevance to vulnerable populations) and “enabling factors” (either individual or organisational characteristics that may influence access to family planning services). The framework was selected due to its past use in prison settings[47] and in identifying key factors that may influence the use of contraception in marginalised communities[48] .

### Study Setting & Description of Current Contraception Policies

In 2023, 768 adult women were held in custody in Australia. Nearly 50% were sentenced (49.9%) and 41.0% were Aboriginal or Torres Strait Islander[49]. Women held in custody in NSW fall under the care of Corrective Services NSW, with the Justice Health and Forensic Mental Health Service (hereon Justice Health NSW) responsible for the provision of health services. This service fully funds all healthcare services in lieu of the availability of Medicare (Australia’s universal healthcare insurance scheme), as well as the provision of medications and commodities (such as the contraceptive pill and IUDs), which would normally be available through the Pharmaceutical Benefits Scheme (PBS), neither of which are available to incarcerated persons [50].

Justice Health NSW policies state that all incarcerated persons should have access to equivalent services that they would be able to receive in the community. This includes the provision or replacement of contraceptive devices, including the oral contraceptive pill, emergency contraception, injectables, implants and IUDs[51]. Aboriginal and Torres Strait Islander women should be offered these services by an Aboriginal health worker [51]. Contraception services can be provided by either the women’s health nurse, midwives as part of post-partum care or a general practitioner (GP). To access any health service, women must submit a written request, which is triaged by nursing staff who will then assign to the appropriate provider pending availability and perceived urgency.

### Participant recruitment

Through a combination of web searches, professional networks, and a snowball sampling approach, we identified staff employed by either Justice Health NSW, community-based organisations involved in providing health services for incarcerated or newly released women or involved in the delivery of family planning services to the general community (n=12). Potential participants were contacted by email with an information sheet about the study, and upon indicating interest in the study, and having questions answered, were asked to provide verbal informed consent prior to participation in the study. Following consent, participants were requested to forward the study information onto other potentially interested colleagues.

### Data collection

Each participant was interviewed for approximately 1 hour on Zoom/Teams and guided by a semi-structured interview schedule which focused on organisational and individual priorities, services provided, demand for services, barriers to service delivery, individual patient characteristics that impact service delivery, and personal views around incarcerated women’s access to reproductive health services. All factors were explored in the context of the provision of contraception, termination of pregnancy, and fertility treatment support.

### Data Analysis

After the data were collected, recordings were transcribed and uploaded to NVIVO12 (QSR, Australia) for analysis. Data was indexed by themes as well as by demographic information including the job description of each participant. Using both inductive and deductive approaches, a thematic framework was developed to identify key choices, challenges and relational conditions required for women to have their reproductive healthcare needs met. As part of the analysis, a 6-phase protocol for thematic analysis (informed by Braun & Clarke) [52] was applied.

Following analysis, the themes identified and data supporting the themes were reviewed by two additional members of the study team (MW and PS) to consider the suitability of the findings as well as individual biases in the interpretation of the results. Furthermore, a draft version of the findings was shared with participants who had indicated they would like to review sensitive information provided. To ensure confidentiality due to the small sample size, participants were labelled into broad categories of GP, nurse and non-government organisation (NGO) worker.

### Ethical considerations

This study was approved by the University of New South Wales Health & Medical Research Committee (#HC210826), Justice Health & Forensic Medicine Network Health & Medical Research (#G1044/21) & Aboriginal Health & Medical Research Council (#1947/22).

## Results

### Participant flow and description of participants

Thirty-two potential participants were contacted via email, of those, three (9%) never responded, 10 (31%) felt they did not meet the eligibility criteria, seven (22%), whilst eligible, were either too busy or did not want to participate, and 12 (38%) consented to an interview.

Seven participants worked for Justice Health NSW and five worked with NGOs. The NGOs represented a mix of providers of family planning services for the general community, an Aboriginal community-controlled health organisation, an advocacy group and case-worker support services for justice-involved women.

Half of the participants (n=6) were GPs, two (16%) were nurses employed in either a midwife or women’s health role, with the remainder having non-clinical roles including advocacy, casework and clinic management. All participants had been involved in the care of justice-involved women for at least five years, two participants identified as Aboriginal and/or Torres Strait Islander, and one participant had previously been incarcerated. None of the participants were attached to a specific prison but rather worked across several facilities or supported women who had come from a variety of locations across NSW.

### Findings

Seven themes were identified and are presented here according to the three main fields of enquiry as defined by the Vulnerable Populations Behavioural Model. They include: 1) predisposing characteristics of incarcerated women, 2) need factors for incarcerated women, and 3) structural factors that influence access to contraception.

We present these themes and their consequences on the outcome of interest, access to contraception, in Figure 1 below.

**Figure 1.**
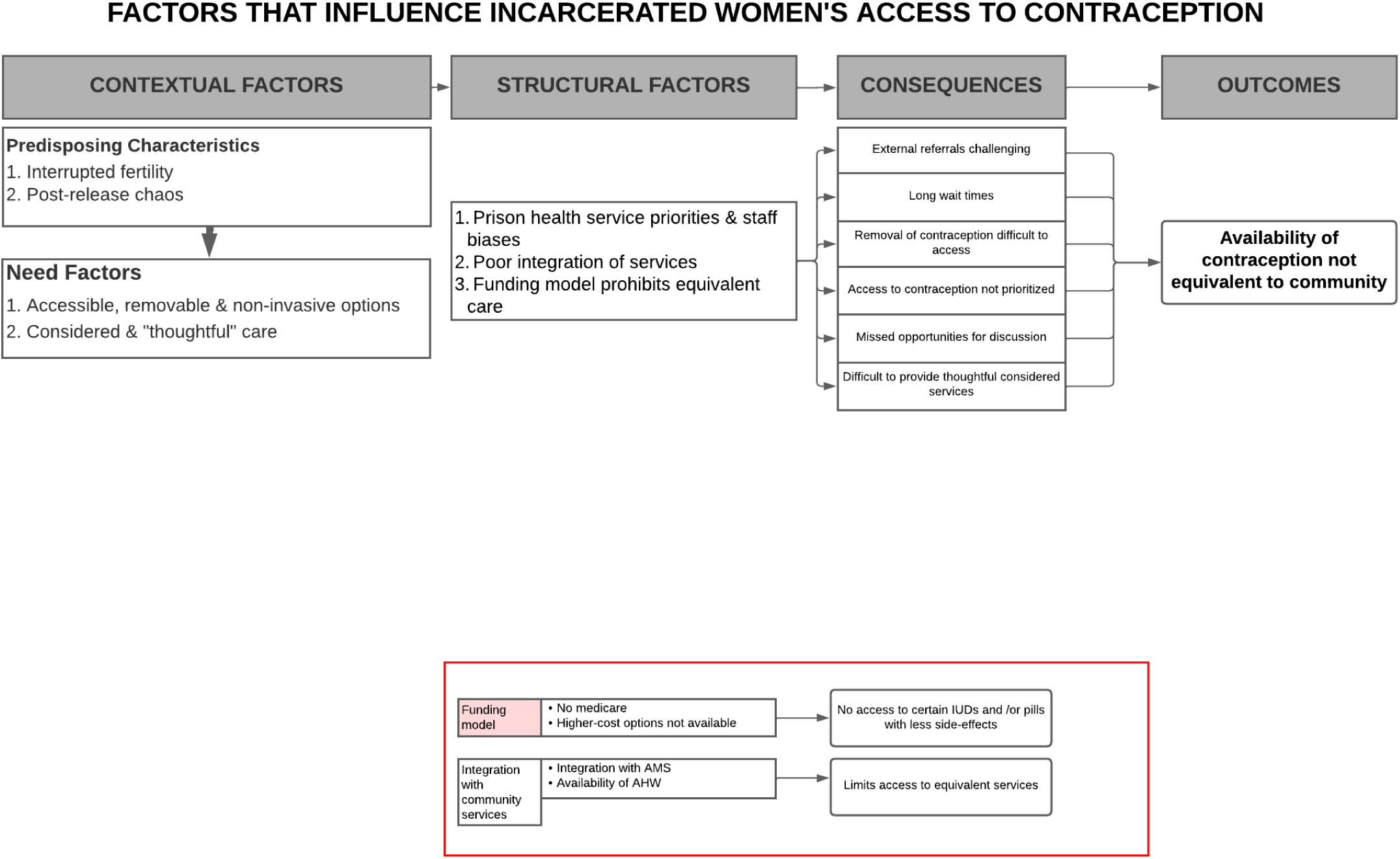
Factors that influence incarcerated women’s access to contraception.

### Predisposing characteristics of incarcerated women

#### Interrupted fertility

Many participants observed that women’s desires around future fertility often became heightened during their incarceration.

> *“What struck me a lot is how frustrated women who’ve been in prison can feel in terms of their fertility…”*

> GP

Staff from both Justice Health NSW and NGOs believed that several factors were driving this sense of interrupted fertility: 1) the nature of incarceration itself halted fertility ambitions; 2) the loss of partners who had then gone on to have children with new partners during their incarceration; 3) menstruation becoming become irregular or stopping completely during incarceration; and 4) correctional service policies that forbade women accessing artificial reproductive technologies such as egg freezing.

Several clinical staff believed that the nature of the prison environment created a heightened anxiety amongst women regarding their fertility opportunities post-release. One GP described these feelings below in the context of women’s periods stopping during their incarceration.

> *“I see a lot of women whose periods have changed when they’ve come into prison, and often they’ll stop. And so, for some people, you know, that can sometimes feel like a convenient thing, but for most of the women, they find it quite a fearsome thing, that they no longer have their periods. And they can see that as a sign of potentially being damaged by the prison environment and their fertility for the future being damaged.”*

> GP

#### Post-release chaos

Participants who worked with women after release noted that this period was often characterised by chaos and uncertainty, fuelled by unstable housing, substance use issues and/or returning to violent relationships.

> *“The chaos of, especially that kind of initial post release period, in terms of relationships, in terms of drug use, in terms of homelessness, and a lot of women kind of, you know, sort of having kind of housing in a sex for favour type sort of set up, like so much”*

> NGO STAFF

They observed that this chaos often caused women to struggle to engage with community health services at release, making it difficult to access contraception.

> *“So, there’s other challenges, the mental health, the communication, you know, unable to communicate their needs… And then, you know, because we’ve got a lot of women that are barred from services as well. So, but that’s, you know, as a result of not getting their needs met, not being able to communicate their needs, and then just sort of giving up because they’re like, nah, I can’t cope with this”*.

> NGO STAFF

NGO staff stated that they often witnessed the consequences of not being able to access contraception in the community, namely demand for support in accessing pregnancy termination services. NGO staff noted that they also often had to support women in need of late term termination services, which were more expensive and harder to access in prison. NGO staff noted that the observed reasons for not being able to access timely abortion services were similar to the reasons they were unable to access contraception, including financial issues, being turned away at the clinic, or feeling judged or stigmatised around their choices.

> *“So sometimes they will go to [THE CLINIC], to, to explore their options and say this is what they want to do. And they don’t feel that people are actually hearing them when they say they want a termination, and they don’t want to have this child. So, they’re like, ‘No, it’s a big decision. You go back and think about it and come back’… and they don’t come back”*.

> NGO STAFF

For NGO staff, this high demand for termination services in the post-release period was indicative of an unmet need for contraception amongst newly released incarcerated women.

> *“If someone is wanting contraception, then the ideal place to have that done, if it’s their choice, is in custody before they are released. So then, when it’s done and dusted, it’s happening, it’s in place. Because once they get out […], it’s really hard”*

> NGO STAFF

### Need characteristics of incarcerated women

#### Accessible, reversible and non-invasive options

Participants observed a high demand for non-invasive and flexible methods including non-permanent, controllable contraception such as Intra-Uterine Devices such as the Mirena^®,^ and rods such as Implanon^®^), short-term injectables such as Depo-Provera® (‘Depo shots’) or the contraceptive pill. They also noted there was often considerable demand for the removal of both IUDs and rods.

While most participants, both NGO workers and clinical staff, believed that hormonal or non-hormonal IUDs may be more suitable given many women’s chaotic lifestyles, they acknowledged that this was not as popular among the women they worked with, who often wanted more flexible and less invasive methods.

> *“We encourage them to go for long-term contraception rather than, you know, adhering to a pill, anything that requires a regime or rather you being adherent to it.”*

> GP

> *“They* [WOMEN] *like the idea of taking a pill that they can stop and start themselves and don’t want that lack of autonomy and, you know, having something you have to have taken out by a doctor, and just access sometimes, it’s just the issue, you know, you’d like an IUD, but there’s no one to take it out.”*

> GP

Depo shots were in particularly high demand, despite both staff and NGO workers feeling that these might not be the most logistically appropriate for women leading chaotic lives as it required regular engagement with a health service. Despite this reservation, they attributed its popularity to being less invasive and short-acting.

> *“There’s a lot of depo, which is what I’ve seen too with women who are coming through drug health services, they would put them on the depo. But like it’s, it’s chaotic for those women as well, because they’ve got to come back for an injection every 12 weeks.”*

> Nurse

#### Considered and “thoughtful” care

All participants recognised that a key component of providing appropriate contraception to women during their incarceration included thoughtful counselling, where women’s individual fertility aspirations post-release and their past experiences needed to be considered. Women with sexual assault histories often needed additional counselling when considering any sort of vaginal insertion, both regarding whether they felt comfortable with this type of method and preparing them for the insertion procedure.

> *“We’re talking about a cohort of women who have generally had very high experiences of sexualized violence, domestic violence, childhood sexual assault […] They’ve been stripped, searched on a regular basis […]. People are in kind of that trauma response [..] And so going through a process of having something like that [VAGINALLY INSERTED] can be actually quite terrifying.”*

> NGO STAFF

> *“So do you know, if you’ve been sexually assaulted, maybe kind of like, you’re not actually that comfortable with people putting things inside your vagina…”*

> GP

Clinical staff also highlighted the importance of being able to refer incarcerated women to specialist services in the community for complex insertions and removals (needed due to anatomical issues, high levels of distress or predisposing medical issues such as epilepsy).

> *“I am not going to put a Mirena® in someone who’s going to jump off the bed on me. Sometimes the women are a bit like that […] I just don’t want to do harm. So, on, you know, if I don’t think that they’re suitable to have it done with me in the jail, then I will send them out.”*

> Nurse

Furthermore, considerations around who should provide the counselling or services, for example, an Aboriginal health worker for Aboriginal and/or Torres Strait Islander women, also needed to be considered.

> *“I’ve heard [INCARCERATED] women talk about […] if they have a First Nations person there and an older First Nations woman there, who’s speaking to them, who’s from outside and just from the community, there is less fear of judgement, there is less fear of discrimination. And there’s less, there is more sense of this person will be okay, if I share this, they’re not going to have a viewpoint of me, this isn’t coming from a practice of trying to annihilate me, disappear me. Judge me, view me in a different way. Because of who I am, aware of my cultural connection.”*

> NGO STAFF

Nearly all staff recognised that their ability to implement such thoughtful and considered counselling was challenging to deliver in the context of where they worked. The factors that contribute to this are discussed under *Structural Factors that Influence Access* below.

### Structural Factors that Influence Access

#### Prison health service priorities & staff biases

Most participants believed that the correctional health system was designed and funded to focus on acute and complex needs. This included a focus on severe mental health problems, substance use issues, and infectious diseases treatment. The consequence being that health issues such as contraception support often fell through the cracks.

> *“People with complex needs end up getting looked after, but maybe the less complex ones might, you know, have less priority and less involvement,”*

> GP

Whilst several Justice Health NSW staff felt that there had been vast improvements to preventative screening services such as mammograms and Pap-smears, several staff reflected that there did appear to be a blind spot amongst some staff when providing comprehensive contraceptive services to women. One nurse described other nurses being genuinely bemused as to why contraception should be prioritised at all.

> *“I think that lack of support from other staff is very frustrating […] We’re trying to do what’s best for them, so that they can get on with their life in the community. I mean, sometimes they say, ‘Oh, why? Why are you doing contraception? […] they don’t need it in jail’. And then we have to explain…. Well, we’re doing this for when they’re released.”*

> Nurse

Several NGO staff suggested that this attitude, in part, related to prioritising immediate health needs rather than considering needs within a continuum of care, both during and after a woman’s incarceration. They also suggested it was due to a de-gendering of women during their incarceration.

> *“Corrective Services has a viewpoint that their responsibility is to keep people safe and contained for the duration of their time […] ‘This is our responsibility. Our responsibility isn’t after. So, this is all I’m going to look out for’. So that is one reason I think that it’s totally missed, because they don’t see it as their responsibility.”*

> NGO STAFF

> *“People aren’t thought about so much in terms of whether or not they want to have babies or whether or not they want to have contraception, they’re thought about as a prisoner […] a kind of minimising of those other aspects of women’s lives when somebody goes to prison […] it’s [REPRODUCTIVE HEALTH] so far down the list in terms of how people are thinking about women in prison, and how people are thinking about access more broadly, and how people are kind of humanising or dehumanising the women that end up in in prison.”*

> NGO STAFF

As a result, most participants agreed that women often experienced challenges accessing appropriate contraception services. This included difficulty arranging external services for complex inserts and removals, long-wait times to meet with the women’s health nurse and most notably, challenges in having contraception removed. According to several participants, factors impacting contraception removal included - the lack of prioritisation of the service, and the perceived “appropriateness” of a woman to be able to have children by staff.

> *“It’s always worried me a bit, because, you know, women will sort of come in and, you know, you can see they’ve got long acting, contraception already in, they’ll tell you, they want it out. Because they want to fall pregnant when they leave prison. But, you know, you can see that, in fact that they’re in a chaotic state stage, and probably not making a good choice to have the contraception out […] Nevertheless, of course, it does become their choice, but I wonder whether providers might be declining to take them out?[…] [WOMEN ARE] feeling unsupported in wanting to have it taken out”*

> GP

#### Poor integration of services

Staff involved in the provision of contraceptive services recognised that that their ability to provide thoughtful and considered counselling around contraception was hampered by structural factors such as a siloed health delivery model, poor integration with community services, particularly in relation to Aboriginal health services, and weak pre-release planning systems.

Services were often provided as targeted interventions rather than an integrated system, which staff noted resulted in lost opportunities for discussions around reproductive health. Several GPs noted that this siloed structure was also creating backlogs for the women’s health nurse, of which there was only one. They described any “female-focused service” being referred to her, creating unnecessarily long waiting lists and pressure on the women’s health nurse to quickly turn around patients, encumbering her ability to have detailed conversations with patients.

> *“It’s all* [REQUEST FOR CONTRACEPTION] *devolved to the women’s health nurse. We’ve only got, I think, one women’s health nurse to the whole of New South Wales. So, you know, she’s absolutely marvellous, but obviously only human, so she’s not going to be able to capture everyone.”*

> GP

Poor integration of family planning services at release meant that opportunities for thoughtful and considered discussion were often missed. This was driven by myriad challenges that impacted all pre-release planning including difficulties to predict departures, chaotic systems and poorly completed paperwork. One GP noted that the extent of these issues varied from centre to centre and referrals related to contraception did take place in smaller facilities, with smaller staff to incarcerated women ratios, combined with a more stable population allowed for more thoughtful planning.

> *“It depends so much on the context. So, I used to work at a smaller prison, which is now at least temporarily closed, which was minimum security and had a strong focus on preparing people for the community. And it really was quite a routine thing for the primary care nursing staff to find out whether, you know, at release, contraception was required, but now, for example, I work in a Remand Centre, which is obviously much more chaotic, with some people released from there and some people in there for a long time.[…] I can well imagine that many women are released without having had that conversation”*

> GP

NGO staff noted that their observations of some women leaving prison without contraception, falling pregnant and then requiring support to access termination services, suggested poor access to contraception as part of pre-release planning.

> *“I don’t think it is something that is prepared for on release […] women do not come out with contraception. So, it’s definitely an area that’s missed.”*

> NGO STAFF

Several participants described a chronic lack of Aboriginal health workers, who were perceived as critical players in providing culturally appropriate care.

> *“They don’t have enough of an Aboriginal workforce that you can just see an Aboriginal person when you want. Yeah, it’s a big, big issue”*

> GP

Aboriginal workforce shortages were generally attributed to ongoing challenges in making Justice Health NSW roles attractive to Aboriginal healthcare providers, combined with poor integration with community services.

> *“There are always open positions for Aboriginal health practitioners, they [ARE] constantly attempting to recruit staff to those positions, and there’s a big turnover… There are many reasons for that. […] I think that a lot of [Aboriginal] people with that sort of qualification are probably quite keen to work in their communities”*

> GP

#### Funding model prohibits equivalent care

Most participants recognised that the suite of contraception types available through the correctional health system was not comparable to what was available in the community. For example, both copper IUDs and certain brands of contraceptive pill (that offered less side-effects) were generally not available due to their higher costs.

> *“We are missing out on the copper IUDs, because I think they’re too expensive for the jail”*

> GP

> *“You’re actually constrained by what I think might be an appropriate pill […]. I do a lot of, like a very thoughtful you know, considered approach, based on the woman and her circumstances. So, some of the pills that I like aren’t available. They would be if we could access the PBS”*

> Nurse

Furthermore, participants noted that the lack of access to services and medicines funded by both Medicare and the PBS, impacted their ability provide equivalent services to what was available in the community. This was particularly noted when providing specific Aboriginal health services.

For example, the funding arrangement limited opportunities for local Aboriginal Medical Services to provide services that were available in the community. One Aboriginal participant noted that if the systems were truly equivalent, community-based Aboriginal health services would be better empowered to provide a comprehensive continuum of care to incarcerated women, allowing for more personalised considerations around reproductive and contraception choices.

> *“We [AMSs] should be resourced to be able to keep in contact with our women. If they get transferred from say regional or remote area into the incarcerated centres within an urban area, then we could communicate with a local AMS that this this lady has been incarcerated, and we need the AMS in that location to be able to go and provide ongoing care… but there’s no funding mechanism to do that”*

> NGO STAFF

Several NGO staff believed that the solution lay in the provision of more innovative and flexible ways of providers being able to claim Medicare rebates whilst providing services to inmates.

NGO participants also suggested that inadequate funding for AMSs to provide justice-based services was in part due to a lack of formal mandate, but also a general lack of interest from AMSs in supporting incarcerated people in the community.

> *“Ya know, for our people, especially in the AMS sector lane, we talk about working with our people from life to death, right? But when they go into jail, we don’t go anywhere near them… I don’t think there’s a major focus on it. And that’s scary.”*

> NGO STAFF

## Discussion

This study is the first in Australia to consider factors that support or impede the provision of contraceptive support to justice-involved women during incarceration and following release. Issues identified, such as health service provider biases and provider reluctance to perform IUD removals mirror issues identified in the community [41–43], they appear to be magnified in a secure setting where individual autonomy and personal self-efficacy are severely limited due to the structural constraints of the prison health-system. Furthermore, unique constraints, created by a funding model which operates outside state funding models and focused on acute healthcare needs, appears to impede women’s ability to access community equivalent contraception services. Given the highly sensitive needs of many of the women who are under the care of Justice Health NSW during their incarceration, characterised by histories of trauma, chaotic lifestyles, and nuanced fertility intentions, our study suggests that far more consideration is required regarding appropriate contraceptive support, if Reproductive Justice is to be achieved for these women.

Several factors were identified requiring further consideration. Firstly, ‘*Structural Factors that Influence Access’* highlighted a focus on acute care needs, including substance abuse, infectious disease and mental health, which whilst important and needed, can result in a lack of resources available to support routine, holistic and patient-focused care during incarceration. The implications of this, such as the emergence of medical emergencies that could have been prevented if given attention earlier, has been extensively reported[34, 53, 54], its consequences on services such as contraception however, have received less attention. Whilst prison-health surveys demonstrate the need for focusing on such acute needs[32, 55], our findings demonstrate the importance of ensuring that the provision of these services do not come at the expense of more future-focused planning of services that may prove to be highly valuable to women, particularly in the post-release period.

Secondly, whilst certain female specific services, such as mammograms, Pap-smear services and antenatal care were reported to be well supported, our theme related to *‘service priorities & staff biases’* highlighted that several participants expressed concerns around the ‘de-gendering’ of women’s needs during incarceration, which was reflected with limited staffing numbers available to deal with women’s health issues. Whilst a sizable body of literature has raised concerns around this problem, this has been examined more within the context of female prison services generally being based on models designed to support men, due to their much higher numbers within carceral settings[56–58]. Our findings suggest a deeper de-humanising issue, where any sense of women being considered as fully formed human beings, with intentions and hopes that go beyond their current incarceration, is removed. Whilst this is certainly also the case for incarcerated men, for women, such hopes can include future pregnancy intentions, or the desire to avoid pregnancy. Ignoring this facet of women’s lives makes it impossible to abide by Reproductive Justice principles in this setting.

Thirdly, our theme *‘funding model prohibits equivalent care’* suggest that the equivalence of care principle[59] is not being upheld regarding the provision of contraception for incarcerated women. Incarcerated women are unable to access the same range of contraception options available through the PBS, nor are they able to access integrated models of care that have been developed in the community in Medicare funded services. This is particularly important for the care of Aboriginal and Torres Strait Islander women who are unable to access health services from the community that have been modelled on culturally safe practices and are delivered and informed by Aboriginal community-controlled healthcare principles and delivery models.

Fourthly, our theme *‘service priorities & staff biases’,* highlighted healthcare provider perceptions around women’s reproductive intentions may be impeding women’s ability to have devices removed. Whilst nearly all participants in this study expressed desires to ensure that women in their care were able to express individual choice and agency in their reproductive desires, they acknowledged that these views could be complicated. Participants acknowledged that concerns around a woman’s ability to support a child, given complex histories involving drug use, homelessness, mental health and/or previous child removals, could lead a provider to create structural barriers to the removal of devices. In an environment where individual agency is minimised, such actions essentially result in completely blocking access to a particular service. Women who are perceived by society as ‘unfit reproducers’, due to economic poverty, social status, (mental) health status, race, and/or life history have historically been at risk of reproductive coercion and structural sterilisations[19, 60]. The nature of many women’s profiles during incarceration makes them particularly vulnerable to approaches that encourage these oppressions either consciously or subconsciously. Whilst forced sterilisation, particularly in the US prison systems have received high levels of attention and criticism[61–63], more subtle forms of reproductive coercion, resulting in women being unable to enact their contraceptive choices, including contraception removal, clearly remain. These require thoughtful consideration with the carceral setting.

Findings from this study have several implications. Firstly, the anecdotal implication that women are leaving prison with a high un-met need for contraception, as suggested by several stakeholder participants, needs to be quantified. Secondly, the findings as outlined here, which represents the views of service providers, need to be triangulated with the views of incarcerated women. Both are currently being explored as part of the 2^nd^ Sexual Health and Attitudes of Australian Prisoners (SHAAP-2) study which is currently being implemented in NSW. Thirdly, these findings provide additional justification as to the importance of incarcerated persons being able to access Medicare services. Finally, as the rate of female incarceration continues to increase both globally [64] and in Australia[31], our study highlights the importance of ensuring that gender-specific services, such as contraception, are provided in a thoughtful and empowering way, as well as gender-appropriate training for both staff and incarcerated women. Whilst there is increasing recognition that key concepts of reproductive justice, such as individual agency in managing reproductive choices, is fundamentally irreconcilable with existing in a state of incarceration[65, 66], attempts must be made to improve the mechanisms by which these services are delivered.

Our findings should be considered in light of several study limitations. Our sample comprised of a small sample size of 12, however, the limited attention that contraceptive services were reported as receiving in the carceral setting, reflects that only a small number of health-care workers are involved in the provision of these services. Our extensive snow-balling strategy indicates that the primary players involved in the provision of contraception were included and, furthermore, the inclusion of NGO staff, who support women outside of Justice-Health gave strength to the perspectives provided by the small sample. However, we should note that we observed two key groups, whose perspectives are not included. Firstly, despite numerous outreach efforts, we received high rates of refusal to participate from nursing staff unless they had a specific reproductive health role. This speaks to the challenges discussed by individuals who did particulate in having contraception recognised as a standard health-care services rather than a specialised service. Secondly, due to needing specific ethical approval from Corrective Services NSW, and the associated delays to the study that this would have caused, we were unable to interview anyone working directly for this organisation. Whilst the delivery of healthcare services is provided by Justice Health NSW, the policy and structural role played by Corrective Services NSW would have been a welcome addition. However, given it took over one year to obtain approvals from Corrective Services NSW, this would have further delayed the study, and speaks to the many challenges of conducting research in the carceral system. Finally, whilst many facets of this study will have replicability in other settings, it is important to note that all participants worked in NSW and for publicly funded prisons. Privately managed prisons may have unique features which merit further exploration.

## Conclusion

The availability of comprehensive and patient-centred family planning services is a human right. Not providing such services represent a missed opportunity in supporting women in the justice system who have distinct and unique needs that may become exacerbated immediately after release. Our findings highlight the complexities of being able to provide the services within a carceral setting, despite the best intentions of staff. Key structural factors that need to be considered, to provide appropriate contraceptive services, include considerations around service priorities, addressing staff biases, better integration of contraception services into other services and better funding models to support equivalence of care principles.

## Data Availability

We are unable to make the data available publicly for a number for reasons related to the ethical restrictions placed by our ethics research committee when providing consent for this study. i. The data set contains highly sensitive information about participants» views of their employers in regards to highly contentious issues relating to access to sexual and reproductive health rights of incarcerated women. Furthermore, the number of staff who work in this area are small - making the likelihood of identification high. Given these concerns, we do not believe the risk of disclosure to be acceptable by making such a dataset publicly available. ii. As part of the consent process, participants were assured that their data would remain private and only be made available with the express permission of the investigator team where we could ensure the ongoing privacy of the data. We therefore believe that making this data (even if de-identified) publicly available would be a breach of our agreement with participants. We are, however, able to provide the data set for review by individuals at PLOS ONE upon request. The contact details for the ethics committee are listed below: NSW Human Research Ethics Committee HREC Ref: # HC210826 02 9348 1943

